# Evoked connectivity of cortical stimulation for memory

**DOI:** 10.64898/2026.01.10.26343861

**Authors:** Eric R. Cole, Riley DeHaan, Lou T. Blanpain, Nealen G. Laxpati, John J. Sakon, Sameer A. Sheth, Kareem Zaghloul, Barbara C. Jobst, Michael R. Sperling, Bradley C. Lega, Gregory A. Worrell, Michael J. Kahana, Robert E. Gross

## Abstract

Despite promising results, it remains unclear how to optimally target and personalize closed-loop stimulation to ameliorate deficits in memory and other cognitive functions. We hypothesized that evoked connectivity – the measurement of neural pathway activation using single pulses of electrical stimulation – can guide patient-specific selection of stimulation location and parameters for memory. We characterized brain-wide evoked connectivity profiles of stimulation in memory-related brain regions recorded from 81 patients undergoing intracranial monitoring for epilepsy, showing that greater evoked connectivity between the lateral temporal cortex and the broader memory network (including the mesial temporal lobe, limbic regions and prefrontal cortex) corroborates observations of memory improvement by lateral temporal cortex stimulation. We first found that the lateral temporal cortex, compared to other stimulated regions, evokes the greatest and most distributed connectivity response throughout other memory-related regions. Evoked connectivity in downstream regions is greatest when stimulating a previously identified optimal target for memory improvement, bordering white matter at the rostrocaudal center of the middle temporal gyrus. Evoked connectivity corroborates other biomarkers of memory improvement by closed-loop stimulation, including increased resting-state functional connectivity between stimulated and recorded sites and increased modulation of oscillatory power. These results provide insight into the network mechanisms of stimulation for memory and suggest that evoked connectivity can more broadly predict the functional effects of closed-loop stimulation to prospectively guide targeting and parameter selection.

## 1. Introduction

Memory dysfunction is a major worldwide health challenge, constituting a major economic burden in Alzheimer’s disease and a common comorbidity in many neurological conditions such as traumatic brain injury, epilepsy, and general aging.^1-4^ Approximately 1 in 25 people suffer from episodic memory loss due to disease.^5^ Electrical brain stimulation techniques to improve memory and ameliorate its dysfunction could improve quality-of-life for patients with memory loss and improve the neuroscientific understanding of memory through targeted manipulation of neural circuits.

Our prior work demonstrated a direct electrical brain stimulation paradigm for improving episodic memory.^2,6^ In patients undergoing intracranial seizure monitoring for refractory epilepsy, we designed patient-specific classifiers that successfully predicted the probability of successful memory recall and applied them to prospectively trigger stimulation during periods of poor encoding. Although effective overall in improving patients’ performance in a verbal free recall task, its success was variable across patients and stimulation locations, necessitating the development of biomarkers that predict the effects of stimulation and could therefore guide stimulation adjustment to achieve the best performance in each patient. Additionally, insights gained from the study of brain stimulation for memory could generalize to other disorders that require precise activation of connectivity, including treatment-resistant depression,^7^ obsessive-compulsive disorder,^8^ Parkinson’s disease,^9^ epilepsy,^10^ and more.

Several sources of variability make it challenging to optimize brain stimulation for memory loss and other conditions (reviewed previously^5,11,12^). First, differences can arise across anatomical regions selected as the stimulation target. In particular, macroelectrode stimulation of key memory regions such as the hippocampus and entorhinal cortex^13^ has been shown to impair memory while stimulation of other regions such as lateral temporal cortex (LTC) and amygdala^14^ can improve it. These differences may arise from variability in the precision of anatomical targeting within the given region (e.g. nearer to vs. further from white matter tracts or boundaries between the stimulated target and adjacent structures) or stimulation parameters such as frequency and waveform pattern that may produce different entrainment effects on activity within the memory network. Stimulation timing relative to different memory states may also affect performance, whether defined according to behavior (e.g. encoding vs. retrieval) or neurophysiology (e.g. pre-stimulation theta rhythms). Last, inter-individual variability could require personalization of any of the above features for effective modulation.

Further work has therefore investigated biomarkers that explain variability in the effects of electrical stimulation on memory and can prospectively inform electrode targeting and parameters. Our initial study of closed-loop stimulation associated successful recall improvement with a restoration of spectral biomarkers associated with good encoding states;^6^ therefore, stimulation could be optimized to engage broadband activity throughout memory-related networks. Second, theta-frequency functional connectivity measured between stimulated and other memory-associated sites using intracranial recordings predicted effective stimulation sites for memory improvement.^15^ Last, anatomical proximity of the stimulation target to white matter tracts predicted memory improvement, suggesting a mechanistic role for widespread network engagement by LTC stimulation.^15^

Despite promising predictive power, however, these biomarkers each suffer from several drawbacks that limit their ability to practically guide stimulation target and parameter selection. First, stimulation-driven oscillatory biomarkers are often noisy, featuring low signal-to-noise properties (requiring repeated measurement over many stimulation trials) and overlap with stimulation artifacts in both frequency and time domains, making their measurement practically challenging.^16^ Functional connectivity or white matter proximity could inform the optimal location to stimulate but are insensitive to adjustments of stimulation parameters, such as the electric current amplitude or pulse width – variables that determine shape and size of the electric field and therefore critically influence which regions and white matter pathways are activated by stimulation.^17-19^ Last, strategies to estimate stimulation-driven white matter activation from anatomical imaging also suffer from imprecision, currently requiring modeling assumptions and population-level normalization or standardized brain atlases, limiting their potential to make accurate and detailed patient-specific predictions regarding stimulation effects.^20-22^

Here, we investigate whether evoked connectivity – the estimation of neuroanatomical connectivity using single-pulse stimulation (often referred to as cortico-cortical evoked potential mapping)^23,24^ – can be used to prospectively optimize targeting and parameters of closed-loop stimulation for episodic memory. Toward this end, evoked connectivity has several useful properties. Features of the evoked potential waveform (for example, the latencies of positive and negative deflections) can be reliably linked to activation of specific anatomical circuits, enabling mechanistic interpretability and potential for further hypothesis generation which are often lacking in translational studies.^25,26^ Evoked connectivity is quickly measurable (within seconds) and exhibits high signal-to-noise properties, enabling precise measurement in relatively few trials – a critical benefit that can accommodate the increasingly complex and high-dimensional parameter spaces available in modern stimulation devices.^27^ Additionally, single-pulse stimulation is generally imperceptible and well-tolerated,^28^ and thus it can circumvent harmful or uncomfortable side effects caused by high-frequency stimulation in various regions.^29^ Last, evoked connectivity is stimulation-induced and responds to changes in stimulation location, amplitude, pulse width, and other parameters in the form of a within-patient functional measurement that requires no assumptions about brain anatomy – a property that may prove vital in diseased brains with abnormal or pathologic connectivity. Evoked connectivity may therefore provide an efficient biomarker for patient-specific optimization of brain stimulation for memory and, more generally, other applications that can benefit from precise targeting of anatomical connectivity.

In this study, we use a large-scale retrospective cohort of stimulation experiments to quantify the evoked connectivity profile of electrical stimulation in various memory-related regions, first showing that LTC stimulation engages a more widespread memory network than other stimulation targets. Second, connectivity to downstream regions is greatest when stimulating a previously identified target for memory improvement adjacent to white matter near the anterior-posterior center of the middle temporal gyrus. Last, greater evoked connectivity correlates with previously studied biomarkers of closed-loop stimulation, including increased functional connectivity and stimulation-driven decreases in oscillatory markers of memory. These results suggest that distributed evoked connectivity can mechanistically explain the effectiveness of LTC as a stimulation target for memory and provide a robust biomarker to prospectively optimize stimulation for memory and other cognitive functions.

**Figure 1:**
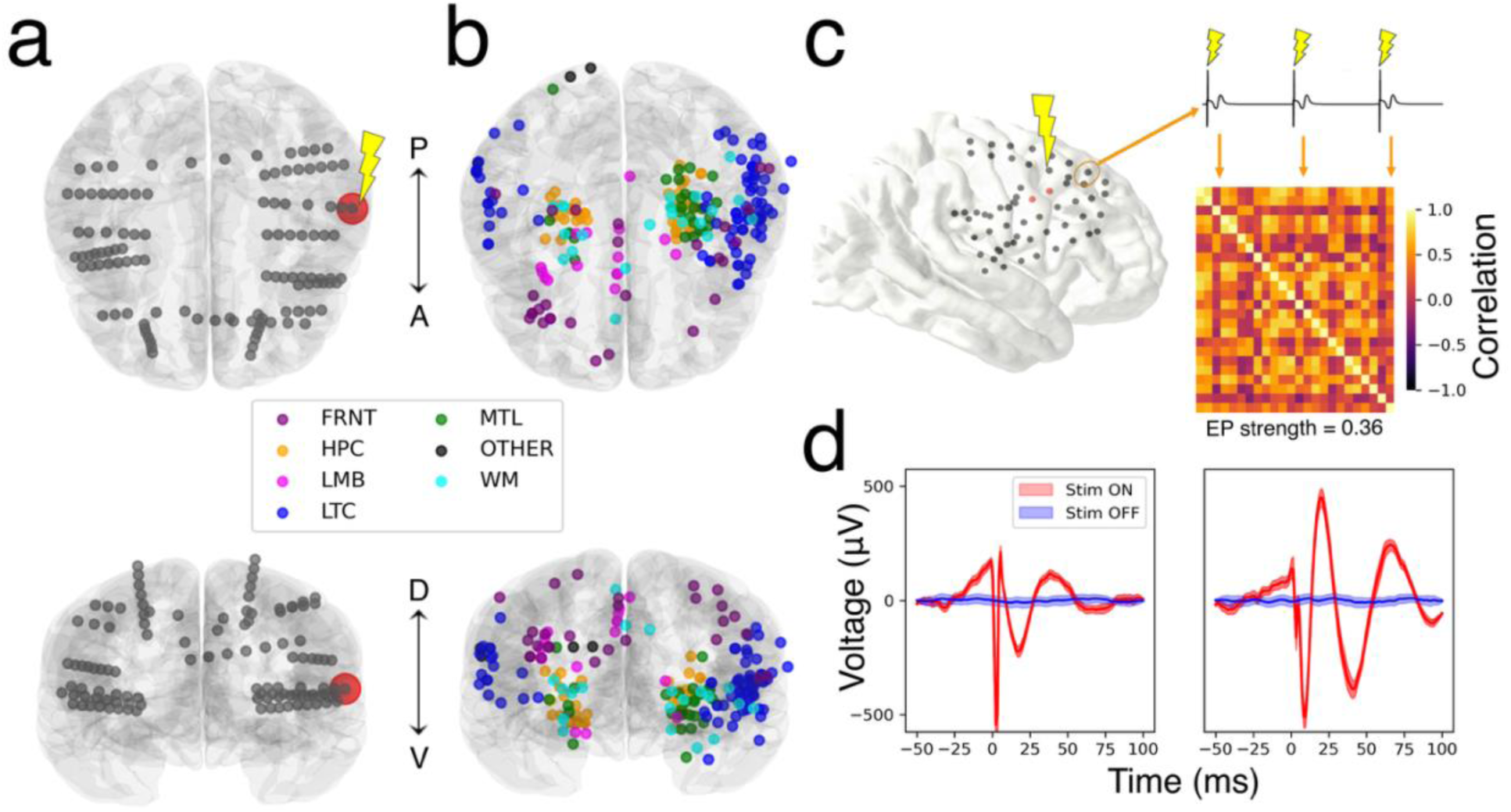
Methods for electrical stimulation and evoked connectivity detection. a) Representative stereoelectroencephalography (sEEG) montage for one example patient. Red: center of a stimulated bipolar pair in lateral temporal cortex. b) All 222 stimulated locations, colored by anatomical location. c) Signal processing for evoked potential detection. For a given recorded site, evoked connectivity is quantified by computing the average time-series correlation between pairs of response waveforms following repeated stimulation pulses delivered at the same site. d) Example channels demonstrating an evoked response (mean +/- standard error across 24 trials). FRNT: prefrontal cortex. HPC: hippocampus. LMB: limbic network. MTL: extrahippocampal mesial temporal lobe. WM: white matter. OTHER denotes contacts in less frequently tested areas (occipital lobe, parietal lobe, subcortical regions, etc). A: Anterior; P: posterior; D: dorsal; V: ventral.

## 2. Methods

### 2.1 Participants

We retrospectively analyzed data collected as part of a multi-institutional effort to evaluate the effect of direct brain stimulation on human memory function (see Mohan et al.^16^ for a detailed experimental protocol). N=81 patients (255 stimulation sites) with refractory epilepsy were surgically implanted with depth, surface grid, and/or surface strip electrodes for seizure localization. Electrode targets and placement were determined by each institution’s clinical team solely based on clinical need. Recordings were conducted across eight medical centers: Columbia University Medical Center (New York, NY), Thomas Jefferson University Hospital (Philadelphia, PA), University of Texas Southwestern Medical Center (Dallas, TX), Emory University Hospital (Atlanta, GA), Dartmouth-Hitchcock Medical Center (Lebanon, NH), Hospital of the University of Pennsylvania (Philadelphia, PA), Mayo Clinic (Rochester, MN), and the National Institutes of Health (Bethesda, MD). All participating patients provided informed consent prior to research under a protocol approved by the University of Pennsylvania institutional review board (IRB). Each participating hospital was sanctioned for research under a reliance agreement with the University of Pennsylvania IRB.

### 2.2 Stimulation parameter search experiment

Experiments were designed to study the brain-wide effects of electrical stimulation applied at different brain regions and parameters. During each session, subjects sat quietly and rested with eyes open as we applied stimulation at various amplitudes and frequencies at a single site. Common stimulation targets included mesial temporal lobe (MTL) and LTC based on ongoing studies examining memory effects of stimulation. A neurologist oversaw all stimulation sessions and performed an amplitude titration procedure to ensure stimulation was safe and determine the maximum amplitude that did not produce afterdischarges or other epileptiform activity.

Stimulation was applied in a bipolar contact configuration, using two neighboring electrode contacts as the anode and cathode to deliver 300 microsecond charge-balanced biphasic rectangular pulses. For each site, we stimulated at frequencies of 10, 25, 50, 100, or 200 Hz, with amplitudes from 0.25 mA up to the site’s maximum allowed amplitude in steps of 0.125 or 0.25 mA. For some subjects, this protocol included interleaved single-pulse stimulation trials. Each trial consisted of stimulation for a 500 ms duration, with a uniformly distributed random delay of 2750-3500 ms between the offset and onset of consecutive stimulation trials. Within each ∼25 minute session that stimulated one location, the order of stimulation trials was randomly shuffled, where each combination of frequency and amplitude was delivered for 24 trials. Individual subjects participated in this stimulation protocol for 1 to 9 different stimulation locations (mean 2.8 sites).

### 2.3 Data processing

#### Evoked connectivity

To quantify evoked connectivity, we examined stimulation trials corresponding to the lowest stimulation frequency (single pulse stimulation, or 10-Hz if single pulse stimulation was not performed) and the closest amplitude to 1 mA (the most common amplitude across all experiments) tested for each patient. We did not study the effect of stimulation amplitude as a covariate of evoked potential effects because the range of tested amplitudes was much lower than in other cortico-cortical evoked potential studies^28^ (because other stimulation parameters were also varied, including frequency). There was no notable difference in amplitudes tested across regions: the median amplitude selected for all regions was 1 mA, except for hippocampus (median 0.75 mA).

We applied a previously validated methodology^30^ to quantify evoked connectivity: first, recording sites were referenced according to a bipolar montage along adjacent contact pairs on each electrode (as previously used for cortico-cortical evoked potential analysis^31,32^). We excluded pairs where one of the contacts overlapped with the stimulated pair from further analysis due to excessive signal corruption by stimulation artifact. For single-pulse stimulation, we extracted the time series between 10-50 milliseconds following each stimulation pulse for a given recording site and stimulation site, then computed a correlation matrix quantifying waveform similarity between all pairs of pulses. The average of this matrix (excluding diagonal elements) is the ‘EP Strength’ metric, which can be interpreted as a coherence value that quantifies the degree to which the evoked response waveform is similar across repeated pulses at one stimulated site. For 10-Hz stimulation trials, we first averaged the waveform across the five individual stimulation pulses within one pulse train (each 100 ms apart), then computed the correlation matrix in the same manner on the averaged waveforms of different stimulation trials and finally averaging over the non-diagonal matrix to provide the EP strength value. While different in implementation, this approach follows similar assumptions and leverages similar properties of the data compared to other recently validated algorithmic approaches for cortico-cortical evoked potential detection.^33^

#### Functional connectivity

We adapted a previously used method for computing resting-state low-frequency functional connectivity (LFC) between channels.^34^ We extracted time series data from non-behavioral countdown periods preceding each trial of a free recall experiment completed by each patient and calculated spectral coherence (normalized cross-spectral density) in the theta-alpha range (5-13 Hz) between each pair of bipolar-referenced channels. This measure reflects the consistency of phase differences between signals at two electrodes, weighted by the correlated change in spectral power at both sites. We used a time-bandwidth product of 4 and a maximum of 8 tapers, computing coherence for frequencies between 5 and 13 Hz. We computed interelectrode coherence within 10 nonoverlapping 1-s windows of data collected during each 10-s baseline countdown period and averaged over windows to derive a single LFC value for each pair of recording sites. For comparison to evoked connectivity, we analyzed the resulting LFC values computed between the stimulated pair and all other recorded locations.

#### Stimulation-driven bandpower changes

To compare evoked connectivity with post-stimulation bandpower changes, we adapted an analysis protocol from prior work.^16,34^ For each pair of stimulation and recording sites, we selected additional stimulation trials from the same experimental session that were performed at 200-Hz stimulation frequency – the frequency that was used during closed-loop memory modulation experiments and most robustly shown to modulate oscillatory biomarkers. We computed multitaper power spectral densities and extracted log-scaled power in theta (4-10 Hz) and high-frequency activity (HFA; 30-100 Hz) ranges at windows of 50-950 milliseconds before stimulation start (pre-stim) and 50-950 ms after stimulation offset (post-stim). To analyze whether evoked connectivity predicts bandpower modulation, we computed the average difference between pre-stim and post-stim bandpower values across trials for each recorded site and compared these values to EP strength for the same stim/recording pair (as derived using low-frequency stimulation trials).

### 2.4 Anatomical localization

As in prior work,the pre-operative structural T1-weighted MRI scan was parcellated, segmented, and coregistered with a post-operative CT scan using Advanced Normalization Tools (ANTS) to provide coordinates. Implanted electrode contacts were manually identified and annotated according to these coordinates. An automated pipeline then mapped these coordinates to brain region labels.^35^ For some subjects, medial temporal lobe subregions were also automatically labelled in a preoperative T2-weighted MRI scan when available.^36^ After obtaining brain region labels, we defined a naming-based protocol to parse automatic region labels into a lower-resolution grouping of hippocampal, lateral temporal, mesial temporal, and other areas of interest for the study. Throughout the remainder of the manuscript, the “memory network” is categorized as this grouping of structures of interest, including the hippocampus, extrahippocampal mesial temporal lobe (including parahippocampal and fusiform gyri), lateral temporal cortex (including temporal gyri and temporal pole), limbic structures (including cingulate gyrus, insula, and amygdala), and frontal lobe (frontal gyri).

### 2.5 Statistical analysis

We used linear mixed-effects (LME) models to examine relationships between physiological variables of interest while modeling the effects of confounding variables, given their ability to account for heterogeneous sampling across groups and conditions. For all EP strength analyses, we computed mean EP strength across trials for each recorded channel and applied the Fisher Z-transform (a conventional statistical approach for converting correlation coefficient values, restricted on a range of -1 to 1, into an approximately normal distribution).^37^ We then accounted for statistical dependence between channels by averaging across channels that were localized to the same brain region for each subject, treating region as the independent unit of analysis, and applying a fixed effect for region category. For example:

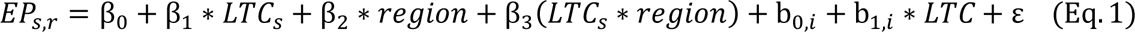

…where EP_s,r_ is the EP strength value for a given stimulated site and recorded region; ‘*LTC_s_*’ indicates whether stim site *s* was localized to LTC; and *b_n,I_* is a random effect coefficient for subject *i*. For analyses comparing different categories of stimulation sites (Fig. 3 and 4), we modeled random effects for the region of the recorded sites (e.g. LTC vs. HPC vs. MTL). and not random effect for subject, as there were few tested stimulation sites per patient (different categories of stimulation sites, for example near vs. far from white matter, were generally not tested within the same subject).

## 3. Results

We first characterize the effects of electrical stimulation in memory-related regions, showing that LTC stimulation evokes the highest average connectivity response throughout the memory network while being among the lowest in the response variance across regions. Second, we examine how anatomical location within the LTC affects evoked connectivity, which is greatest when stimulating near the center of the LTC anterior-posterior axis, adjacent to white matter (particularly when recording from the mesial temporal lobe), and in the middle temporal gyrus. Next, we show that channels featuring a high evoked response to stimulation also display higher resting-state low-frequency functional connectivity (LFC) to the stimulation site and greater modulation of theta-range and high-frequency oscillations in response to stimulation. These results suggest that evoked connectivity is a biomarker that could enable prospective optimization of closed-loop stimulation for memory.

### 3.1 Evoked connectivity of stimulation in memory-related regions

Figure 2 demonstrates the network-wide connectivity profile induced by stimulation in various memory-related regions, including the LTC, hippocampus (HPC), extrahippocampal mesial temporal lobe regions (MTL), frontal lobe (FRNT), and limbic regions (LMB). This connectivity atlas reveals several expected anatomical trends. First, for each region, the highest average response strength was observed locally, i.e. within the stimulated region. LTC stimulation (Fig. 2a) evoked a distributed connectivity profile that was greatest in LTC, frontal, and limbic sites. Hippocampal stimulation (Fig. 2b) evoked a lopsided response profile that was strong within the hippocampus, but comparatively reduced in other regions – however, extrahippocampal MTL and limbic sites were also activated. Stimulation in extrahippocampal MTL regions (Fig. 2c) evoked an overall lower-magnitude response profile that was greatest in MTL, hippocampal, and limbic sites. Prefrontal stimulation (Fig. 2d) evoked a response profile that was highest in frontal, LTC, and limbic regions. Stimulation in limbic regions (Fig. 2e) evoked a response profile that was highest in limbic and prefrontal regions.

**Figure 2:**
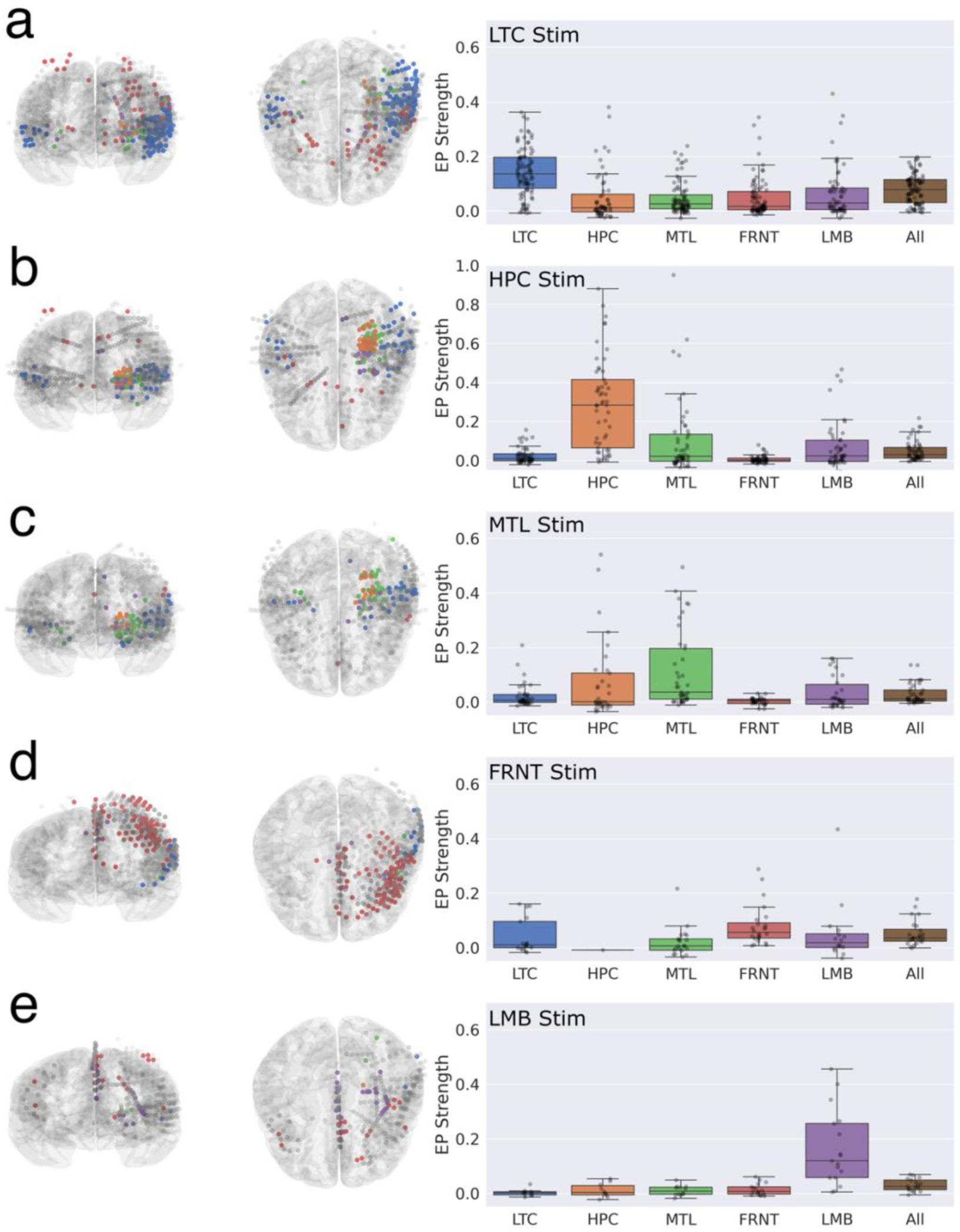
Evoked connectivity of electrical stimulation in memory-related brain regions: comparison across recorded sites. a-e) Left: frontal and dorsal views of evoked responses to stimulation at a given region, aggregated across up to 25 stimulation sites per region. Colored coordinates: channels that featured an above-threshold evoked response to stimulation (EP strength > 0.15) at the given region. Gray coordinates: channels with a below-threshold stimulation response. Right: EP strength resulting from stimulation of the given region, averaged across channels that were localized to the named region category. N = 102 LTC sites; 58 HPC sites; 42 extrahippocampal MTL sites; 26 prefrontal sites; and 18 limbic sites. All: average across all channels localized to any of the other five named regions.

Next, we visualized the same response data by stimulation target to compare side-by-side how each target affects each region’s evoked response strength (Fig. 3). Among all regions stimulated, LTC engaged higher average EP strength throughout the memory network compared to other stimulation sites (Fig. 3f) (p = 0.0037; LME; N = 5606 region/site pairs, 102 LTC vs. 144 other stimulated sites), while featuring comparably low variance in EP strength across different regions (Fig. 3g). This broad connectivity profile between LTC and multiple other regions involved in memory encoding could explain its superiority over other regions as a stimulation target for episodic memory improvement.^6^

**Figure 3:**
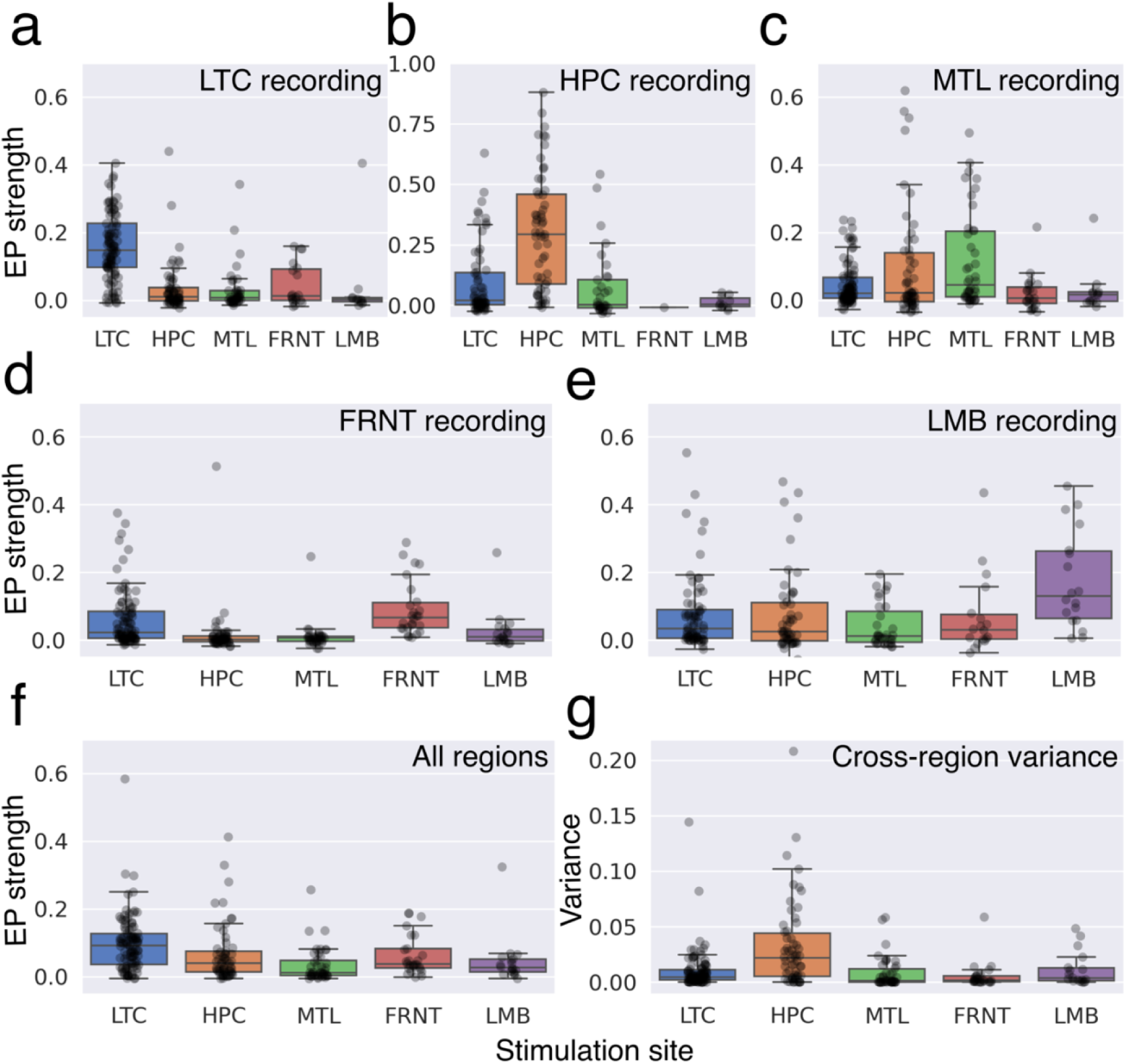
Evoked connectivity of electrical stimulation in memory-related regions: comparison across stimulated sites. a-e) These panels show the same data as Figure 2, transposed to directly compare the effect of different stimulation sites on the same region side-by-side. f) EP strength averaged across all five regions of interest for different stimulation sites. g) Variance of EP strength across different regions, compared between simulation sites.

We subsequently investigated how the evoked connectivity profile varies by anatomical location of the stimulated site within LTC. In particular, we asked whether evoked connectivity explains previously observed anatomical trends of stimulation location for memory improvement, which was greatest when stimulating adjacent to white matter near the anterior-posterior center of the middle temporal gyrus.^6^ Fig. 4a shows the 83 different stimulation sites localized to the LTC, which includes 11 sites localized to the boundary of LTC and white matter. Stimulation in the center of the anterior-posterior axis (middle quintile of stimulation site y-coordinates) produced a higher average evoked connectivity profile in non-LTC memory regions than those in outer segments (Fig. 4b) (p < 0.0001; LME; N = 1321 region/site pairs, 22 inner sites vs. 80 outer sites). Stimulation sites adjacent to white matter evoked greater activation of other regions, particularly the hippocampus and other mesial temporal structures (Fig. 4c) (p = 0.031; LME; N = 1321 region/site pairs, 90 grey matter sites vs. 11 boundary sites). Last, stimulation sites in the middle temporal gyrus evoked a greater average response than in the superior or inferior temporal gyri (Fig. 4d) (p = 0.000045; LME; N = 1267 region/site pairs, 74 MTG vs. 17 ITG, 7 STG sites). Therefore, distributed multi-region engagement by LTC stimulation – most notably in the hippocampus – could explain anatomical trends observed in prior closed-loop stimulation experiments, including the effects of white matter and cortical position.

**Figure 4:**
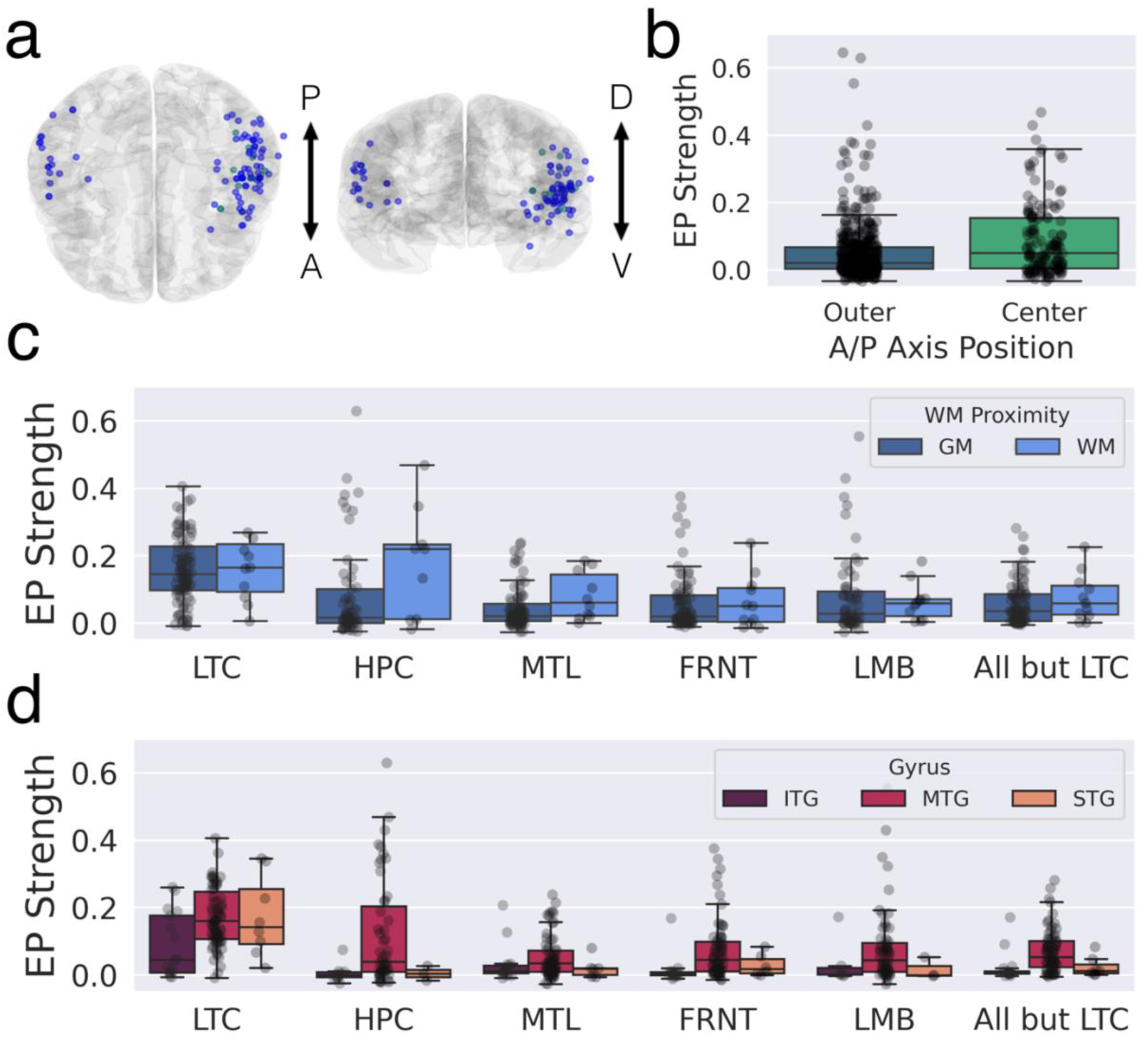
Downstream evoked connectivity is maximized upon stimulating white matter in the rostrocaudal center of middle temporal gyrus. a) Stimulation sites (N=101) localized to the LTC (blue) or boundary of LTC and white matter (turquoise). b) Average EP strength induced in distal regions (mesial temporal, frontal and limbic regions) vs. stimulation location along the LTC anterior-posterior axis. Center: stimulation sites corresponding to the middle quintile of localized stimulation site y-coordinates. Outer: stimulation sites corresponding to stimulation site y-coordinates outside the middle quintile. c) Average EP strength induced in different regions by stimulation sites localized to LTC gray matter only (‘GM’) or to the boundary of LTC gray matter and white matter (‘WM’). d) Average EP strength induced in distal regions by stimulation sites localized to different LTC gyri. ITG: inferior temporal gyrus. MTG: middle temporal gyrus. STG: superior temporal gyrus.

We further investigated whether evoked connectivity can predict physiological biomarkers associated with episodic memory improvement by closed-loop stimulation. We computed LFC^34^ (Fig. 5a) between the stimulated site and all other contacts during the rest period of a free recall task, during which no stimulation was applied. LFC was greatest between the LTC and other LTC sites, followed by MTL and limbic regions (Fig. 5b). Channels with high EP strength also demonstrated higher LFC with the stimulation site (Fig. 5c) (p < 0.00001; LME, N = 6086 region/site pairs, 255 stimulated sites).

**Figure 5:**
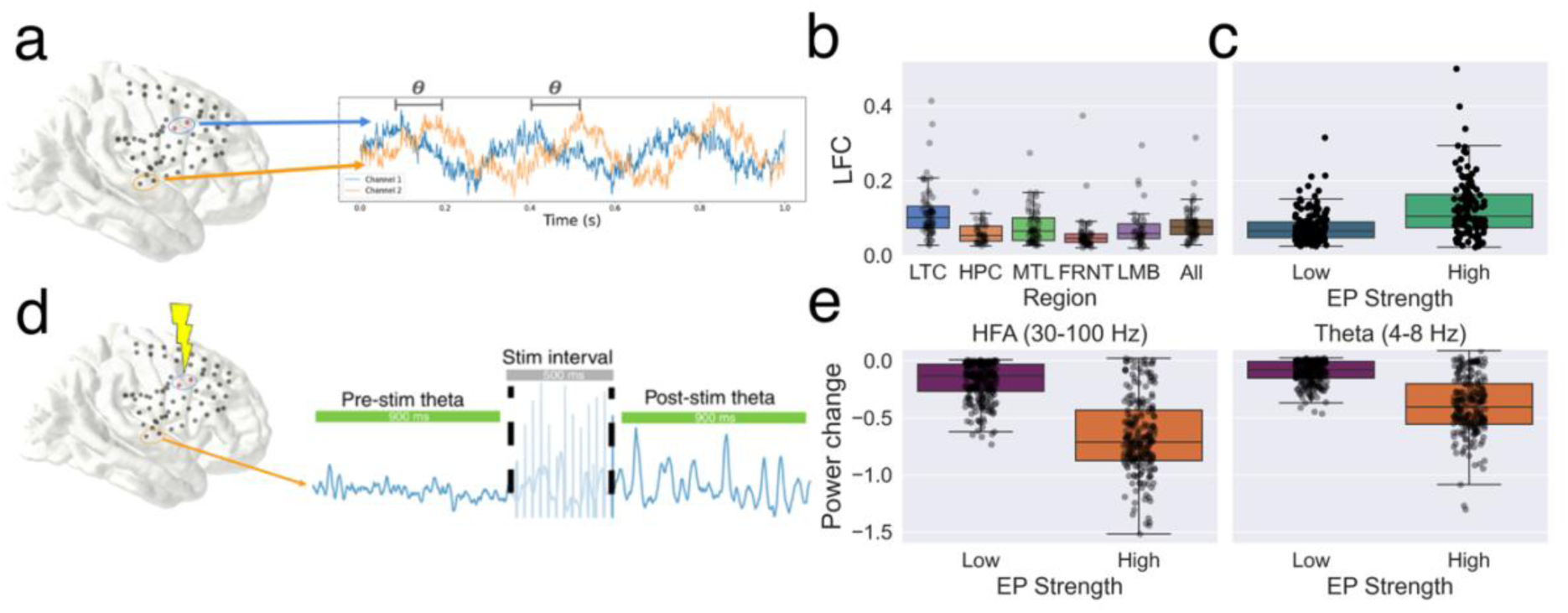
Evoked connectivity predicts increased functional connectivity and modulation of spectral power. a) LFC measures the coherence of low-frequency (5-13Hz) spectral activity between activity recorded from the stimulated bipolar pair (blue circle) during non-stimulated periods and other bipolar recording channels (orange circle). b) Average LFC between LTC stimulation sites and other regions. c) Average LFC between stimulated sites and recorded sites for channels displaying a below-threshold stimulation response (’Low’: EP strength < 0.15) and above-threshold stimulation response (‘High’: EP strength > 0.15). d) Modulation strength is measured by the difference between post-stimulation power and pre-stimulation power at a given channel (orange circle) in response to stimulation (blue circle). e) Average modulation strength of high-frequency power (left) and theta power (right) for channels displaying low EP strength (’Low’: EP strength < 0.15) and high EP strength (‘High’: EP strength > 0.15).

We additionally investigated whether evoked connectivity (measured using low-frequency stimulation) can predict how high-frequency stimulation, at the same site within the same participants, modulates oscillations throughout the memory network. High-frequency stimulation decreased oscillatory power in both high-frequency (p < 0.00001; LME intercept test; N = 6227 region/site pairs, 255 sites) and theta bands (p = 0.00023; LME intercept test; N = 6227 region/site pairs, 255 sites) for low EP strength channels (reflecting our prior study of high-frequency stimulation^16^), and this suppression effect was comparatively greater in high EP strength channels than low EP strength channels for both frequency bands (HFA: p < 0.00001; LME; N = 6227 region/site pairs, 255 sites) (theta power: p = 0.00034; LME; N = 6227 region/site pairs, 255 sites). As spectral power modulation is another predictor of stimulation-driven memory improvement, this result provides further support that evoked connectivity can predict the memory improvement effects of LTC stimulation.

## 4. Discussion

We characterized the effects of electrical stimulation in various memory-related regions, first showing that lateral temporal cortex stimulation engages a more distributed and low-variance connectivity profile throughout the memory network compared to other regions. Downstream connectivity was greatest when stimulating adjacent to white matter near the center of the LTC anterior-poster axis in the middle temporal gyrus, a previously identified hotspot for memory improvement. Channels displaying high evoked response strength also demonstrated higher resting-state LFC and greater inhibition of theta-range and high-frequency oscillations, suggesting more broadly that evoked connectivity mediates the functional effects of electrical stimulation on brain networks. Importantly, this stimulation protocol was not associated with seizures and was well-tolerated by patients.^38^

Notably, the observation that LTC stimulation maximizes distributed connectivity throughout the memory network may partially explain the unexpected finding that LTC is a more effective stimulation target than other regions more conventionally associated with memory, such as the hippocampus and entorhinal cortex. One possible explanation is that the effects of LTC stimulation are limited to types of memory where the LTC may play a more prominent role that relates to its well-characterized involvement in language and audition, such as verbal and language-based memory tasks. It remains unknown whether the memory effects of LTC stimulation would generalize to other modalities, such as visual or spatial memory. A second possibility is that this effect is unrelated to the functional role of LTC and is instead primarily due to its anatomical position. Despite having limited direct anatomical connectivity to mesial temporal areas,^39^ the LTC lies anatomically close to multiple structures of interest, and therefore stimulation at a single site may more easily engage the memory network than other targets. In fact, it remains unclear whether the effects of LTC stimulation on memory result solely from direct activation of white matter tracts passing adjacent to the cortex, activation of LTC grey matter and its axonal projections to other regions, or whether a combination of both is necessary.^40^ The importance of white matter proximity and delineation of a focal optimal target suggest that activation of passing fibers is a plausible, but not necessarily exclusive, mechanism for memory stimulation.

Beyond memory, the association between evoked connectivity and other physiological markers of stimulation suggests that this measure could provide a generalizable biomarker for predicting the functional effects of brain stimulation across networks. From a neuroscientific perspective, this finding supports the concept that network connectivity structure shapes how information or activity propagates throughout the brain in response to perturbations.^34,41^ Similarly, increasing evidence suggests that achieving effective therapy with deep brain stimulation for various neurological and psychiatric applications, such as epilepsy,^42^ Parkinson’s disease,^27,43,44^ and depression^7,45^ can depend critically on selective activation of one or more white matter pathways in or around the region being stimulated. Despite this growing clinical interest, practical approaches for prospectively optimizing stimulation to activate desired neural pathways remain limited. One prominent strategy integrates medical imaging and biophysical modeling to estimate white matter activation, which requires significant computational infrastructure and technical expertise.^20,21^ Other strategies use passively recorded neurophysiological biomarkers to choose a stimulation site or contact arrangement, for example the growing clinical use of beta power in deep brain stimulation for Parkinson’s disease^46^ – a technically convenient method but one that can feature interpretational challenges and limited relevance to the ‘active’ effects of stimulation perturbations and related parameters.

Evoked potential mapping offers complementary advantages and limitations relative to these existing techniques. As we described earlier, evoked connectivity is technically convenient, provides within-patient measurement reflective of ‘active’ stimulation perturbations, and enables high-throughput measurement with high signal-to-noise properties and short measurement timescales – factors that make this biomarker well-suited for patient-specific parameter optimization. Additionally, single-pulse stimulation is generally imperceptible and well-tolerated,^28^ and has shown potential to predict harmful or uncomfortable side effects of high-frequency stimulation without causing them directly.^42,47^ In contrast to image-based methods, evoked connectivity mapping is constrained by the need for neural recording hardware. However, this need is synergistic with the existing clinical use of brain-wide sEEG, which is commonplace for epilepsy, leveraged as a research platform for other conditions, and recently being explored for alternate applications in psychiatry,^48^ chronic pain,^49^ and movement disorders.^50^ Additionally, noninvasive methods such as scalp EEG^51,52^ and electromyography^44,53^ can be used to record evoked potentials and could enable more widespread clinical use.

These results could guide the design of future experiments that prospectively study the relationship between evoked connectivity and memory improvement. In particular, we envision an experiment that leverages the high-throughput potential of evoked connectivity mapping to test a wide range of implanted stimulation sites, amplitudes, and pulse width values throughout the lateral temporal cortex for a given patient. These data could then be used to prospectively determine a smaller set of personalized stimulation settings that maximally engage certain pathway profiles of interest: for example, maximizing connectivity to only LTC, maximizing connectivity to mesial temporal lobe regions without activation of prefrontal and limbic structures, activation of prefrontal and limbic regions without mesial temporal activation, and maximizing connectivity with both sets of pathways. This narrower set of stimulation conditions could then be tested in closed-loop stimulation experiments during a memory task to prospectively evaluate which network activation profile is most mechanistically critical to the memory improvement effects of LTC stimulation (mirroring prior trial designs for neuropsychiatric DBS^54^). Additional work could seek to further understand the physiological relationship between evoked connectivity and brain-wide oscillatory modulation by studying how the temporal and spatial effects of stimulation change as a function of increasing stimulation frequency, duration, and burst energy.^55,56^

This study has several limitations, particularly resulting from the constraints of retrospective analysis of data collected from neurosurgical patients. Our partial reliance on 10-Hz stimulation trials restricted analysis of evoked potential waveforms to a window of less than 100 ms following the stimulation pulse. However, other studies of cortico-cortical evoked potentials have commonly shown response waveforms that also feature decaying response components lasting hundreds of milliseconds,^23^ which could have value in predicting behavior. Recent mechanistic studies suggest that longer-lasting response components are mediated by recurrent cortico-thalamo-cortical network loops,^57^ and therefore our analysis would primarily measure activation of polysynaptic direct cortico-cortical connections.^58^ Additionally, our study did not consider how evoked connectivity varies across subregions within broad area categorizations such as temporal or frontal cortex. This property could similarly confound the comparison of different stimulation sites’ connectivity profiles, as all sites being stimulated or measured downstream of stimulation are unlikely to have, for example, similar proximity to white matter. Similarly, we did not have sufficient coverage of tested stimulation sites to study the effects of specific structures within mesial temporal or limbic groupings that may be important for memory, such as the entorhinal cortex. Last, our evoked potential signal processing methods used only waveform similarity to derive a coherence metric of the stimulation response; thus, we did not examine whether increased evoked response magnitude affects any of the results (though these properties may likely be correlated).

Our proposal of evoked connectivity as a biomarker for memory improvement complements a growing body of research on algorithms for automating stimulation parameter selection and developing closed-loop strategies for cognitive modulation.^27^ This biomarker could be used as feedback for optimization algorithms that can automatically search for optimal settings based on biomarker values (such as Bayesian optimization,^43^ its extensions,^59-61^ and related methods^62-64^) to ensure optimal network engagement for each patient. In particular, the combination of efficient search algorithms with the fast measurement of evoked potentials could practically enable systematic parameter optimization within the time constraints of clinical research experiments, enable practical clinical deployment of cognitive brain stimulation methods, and provide a pathway towards practical use of high-complexity next-generation brain stimulation devices. In the long term, such advancements could improve outcomes for memory loss and other complex conditions by making brain stimulation therapies more precise, reliable, and scalable.

## Data Availability

All data in the present work are available online upon request at:
https://memory.psych.upenn.edu/RAM_Public_Data

## Acknowledgements

This work was supported by the DARPA Restoring Active Memory (RAM) program (Cooperative Agreement N66001-14-2-4032).

